# Persistent clotting protein pathology in Long COVID/ Post-Acute Sequelae of COVID-19 (PASC) is accompanied by increased levels of antiplasmin

**DOI:** 10.1101/2021.05.21.21257578

**Authors:** Etheresia Pretorius, Mare Vlok, Chantelle Venter, Johannes A. Bezuidenhout, Gert Jacobus Laubscher, Janami Steenkamp, Douglas B. Kell

**Affiliations:** Department of Physiological Sciences, Faculty of Science, Stellenbosch University, Stellenbosch, Private Bag X1 Matieland, 7602, South Africa; Central Analytical Facility: Mass Spectrometry Stellenbosch University, Tygerberg Campus, Room 6054, Clinical Building, Francie van Zijl Drive Tygerberg, CAPE TOWN, 7505; Mediclinic Stellenbosch, Stellenbosch 7600, South Africa; PathCare Laboratories, PathCare Business Centre, PathCare Park, Neels Bothma Street, N1 City 7460, South Africa; Department of Biochemistry and Systems Biology, Institute of Systems, Molecular and Integrative Biology, Faculty of Health and Life Sciences, University of Liverpool, Crown St, Liverpool L69 7ZB, UK; The Novo Nordisk Foundation Centre for Biosustainability, Building 220, Chemitorvet 200, Technical University of Denmark, 2800 Kongens Lyngby, Denmark

**Keywords:** COVID-19, Long COVID/PASC, Fibrin(ogen), Microclot, Proteomics, Antiplasmin, Serum Amyloid A

## Abstract

Severe acute respiratory syndrome coronavirus 2 (SARS-Cov-2)-induced infection, the cause of coronavirus disease 2019 (COVID-19), is characterized by acute clinical pathologies, including various coagulopathies that may be accompanied by hypercoagulation and platelet hyperactivation. Recently, a new COVID-19 phenotype has been noted in patients after they have ostensibly recovered from acute COVID-19 symptoms. This new syndrome is commonly termed Long COVID/Post-Acute Sequelae of COVID-19 (PASC). Here we refer to it as Long COVID/PASC. Lingering symptoms persist for as much as 6 months (or longer) after acute infection, where COVID-19 survivors complain of recurring fatigue or muscle weakness, being out of breath, sleep difficulties, and anxiety or depression. Given that blood clots can block microcapillaries and thereby inhibit oxygen exchange, we here investigate if the lingering symptoms that individuals with Long COVID/PASC manifest might be due to the presence of persistent circulating plasma clots that are resistant to fibrinolysis. We use techniques including proteomics and fluorescence microscopy to study plasma samples from healthy individuals, individuals with Type 2 Diabetes Mellitus (T2DM), with acute COVID-19, and those with Long COVID/PASC symptoms. We show that plasma samples from Long COVID/PASC still contain large anomalous (amyloid) deposits. We also show that these anomalous deposits in both acute COVID-19 and Long COVID/PASC plasma samples are resistant to fibrinolysis (compared to plasma from controls and T2DM), even after trypsinisation. After a second trypsinization, the persistent pellet deposits were solubilized. We detected various inflammatory molecules that are substantially increased in both the supernatant and trapped in the solubilized pellet deposits of acute COVID-19 and Long COVID/PASC, versus the equivalent volume of fully digested fluid of the control samples. Of particular interest was a substantial increase in α(2)-antiplasmin (α2AP), various fibrinogen chains, as well as Serum Amyloid A (SAA) that were trapped in the solubilized fibrinolytic-resistant pellet deposits. Clotting pathologies in both acute COVID-19 infection and in Long COVID/PASC might therefore benefit from following a regime of continued anticlotting therapy to support the fibrinolytic system function.

## INTRODUCTION

Severe acute respiratory syndrome coronavirus 2 (SARS-Cov-2)-induced infection, the cause of coronavirus disease 2019 (COVID-19), is characterized by acute clinical pathologies, including various coagulopathies that may result in either bleeding and thrombocytopenia, hypercoagulation, pulmonary intravascular coagulation, microangiopathy venous thromboembolism or arterial thrombosis (Gupta et al., 2020, Perico et al., 2021, Gerotziafas et al., 2020, Siddiqi et al., 2021, Grobbelaar et al., 2021, Grobler et al., 2020, Kell et al., 2020, Pretorius et al., 2020, Venter et al., 2020). Acute COVID-19 infection is also characterized by dysregulated, circulating inflammatory biomarkers, hyperactivated platelets, damaged erythrocytes and substantial deposition of microclots in the lungs (Grobler et al., 2020, Pretorius et al., 2020, Venter et al., 2020, Roberts et al., 2020, Renzi et al., 2020, Ciceri et al., 2020, Bobrova et al., 2020, Lam et al., 2020, Berzuini et al., 2020, Akhter et al., 2020). Acute COVID-19 patients may suffer from thrombocytopenia that may lead to life-threatening disseminated intravascular coagulation (DIC) (Terpos et al., 2020). Predisposing risk factors or co-morbidities that may also lead to a poor prognosis of acute COVID-19, are cardiovascular disease, diabetes, arterial hypertension, obesity (Pretorius et al., 2020, Venter et al., 2020, Finer et al., 2020, Yates et al., 2020, Sattar et al., 2020, Gerotziafas et al., 2020, Li et al., 2020a), as well as cancer (Malek et al., 2020). Complications like liver injury, acute respiratory distress syndrome (ARDS), sepsis, myocardial injury, renal insufficiency and Multiple Organ Dysfunction Syndrome (MODS) are common in cancer patients with COVID-19 (Malek et al., 2020). Plasma of COVID-19 patients also carries a significant load of preformed amyloid clots (Grobler et al., 2020) and this phenomenon may be indicative of a poor prognosis.

Recently, a new COVID-19 phenotype has been noted in patients after they have ostensibly recovered from acute COVID-19 symptoms. This new syndrome is commonly termed Long COVID/ Post-Acute Sequelae of COVID-19 (PASC) (Proal and VanElzakker, 2021). We use the terminology Long COVID/PASC. Long COVID/PASC can involve sequelae and other medical complications that last for weeks to months after initial recovery, and may include more than 50 long-term effects (Lopez-Leon et al., 2021). Preliminary data about Long COVID/PASC symptoms show numerous similarities to chronic illnesses (Carfí et al., 2020, Rubin, 2020, Baig, 2020, Proal and VanElzakker, 2021) known to be associated with viral infections, such as Myalgic Encephalomyelitis/Chronic Fatigue Syndrome (ME/CFS) (Wostyn, 2021, Mohabbat et al., 2020), postural orthostatic tachycardia syndrome(Goldstein, 2021) and Mast Cell Activation Syndrome (Afrin et al., 2020, Huang et al., 2021).

Lingering symptoms have been found to persist for as much as 6 months (or longer) after acute infection, where COVID-19 survivors complain of recurring fatigue or muscle weakness, being out of breath, sleep difficulties, and anxiety or depression (Huang et al., 2021). Patients who were more severely ill during their hospital stay tended to have more severe impaired pulmonary diffusion capacities and abnormal chest imaging manifestations, and are the main target population for interventions for long-term recovery (Huang et al., 2021). However, it was also shown that ∼32% of subjects reporting symptoms at 61+ days after infection were asymptomatic at the time of initial SARS-CoV-2 testing (Huang et al., 2021). Many patients are also developing Long COVID/PASC after mild or asymptomatic infection, despite not being hospitalized (Tabacof et al., 2020). Researchers in Italy found that 87.4% of 143 COVID-19 patients reported at least one symptom 60 days post-infection, and 55% had three or more. According to their research (Carfí et al., 2020), the most common symptoms were: fatigue (53.1%), difficulty in breathing (43.4%), joint pain (27.3%) and chest pain (21.7%). In the UK it was found in a study of 384 patients (mean age 59.9 years; 62% male) followed for a median 54 days post discharge, 53% reported persistent breathlessness, 34% cough and 69% fatigue and 14.6% had depression. In those discharged with elevated biomarkers, 30.1% and 9.5% had persistently elevated D-dimer and C reactive protein, respectively. 38% of chest radiographs remained abnormal with 9% deteriorating(Mandal et al., 2020). In the largest global study to-date on this issue, a survey of 3,762 Long COVID/PASC patients, from 56 countries found nearly half still could not work full-time six months post-infection, due mainly to fatigue, post-exertional malaise, and cognitive dysfunction (Komaroff and Bateman, 2020).

In the current study, we investigate if the lingering symptoms that individuals with Long COVID/PASC might be due to the presence of persistent circulating plasma clots that are resistant to fibrinolysis. We have previously shown that large anomalous (amyloid) fibrin(ogen) deposits are present in plasma from acute COVID-19 patients (Pretorius et al., 2020, Venter et al., 2020). Here we show that plasma samples from Long COVID/PASC still contain large anomalous (amyloid) deposits, and that these deposits are most resistant to fibrinolysis, even with the treatment of a two-step trypsin method. We used proteomics to study the protein presence in both digested supernatant and trapped persistent pellet deposits (after protein digestion via trypsin). Of particular interest was a substantial increase in the acute phase inflammatory molecule Serum Amyloid A (SAA4) and α(2)-antiplasmin (α2AP) that were trapped in the fibrinolytic-resistant pellet deposit. The plasmin-antiplasmin system plays a key role in blood coagulation and fibrinolysis (Schaller and Gerber, 2011). Plasmin and α2AP are primarily responsible for a controlled and regulated dissolution of the fibrin polymers into soluble fragments (Schaller and Gerber, 2011, Miszta et al., 2021).

These results suggest that at least some Long COVID/PASC symptoms may result from persistent and circulating anomalous (amyloid) deposits, together with a pathological fibrinolytic system, that may be related to a substantial increase in certain pathological proteins that interfere with fibrinolysis.

## MATERIALS AND METHODS

### Ethical clearance

Ethical clearance for the study was obtained from the Health Research Ethics Committee(HREC) of Stellenbosch University (South Africa) (reference: N19/03/043, project ID: 9521). The experimental objectives, risks, and details were explained to volunteers and informed consent were obtained prior to blood collection. Strict compliance to ethical guidelines and principles Declaration of Helsinki, South African Guidelines for Good Clinical Practice, and Medical Research Council Ethical Guidelines for Research were kept for the duration of the study and for all research protocols.

### Sample demographics and considerations

Blood was collected from healthy volunteers (N=13; 6 males, 7 females; mean age: 52.4 ± 4.8) to serve as controls. Healthy volunteers did not smoke, or suffer from cardiovascular diseases or coagulopathies and pregnancy, lactation, and the use of anticoagulants, were exclusion criteria (Pretorius et al., 2010). Patients diagnosed with COVID-19 (before treatment) (N=15; 6 males and 5 females; mean age: 54.8 ± 15.3) and also patients with Type 2 Diabetes Mellitus (T2DM) (N=10; 7 males and 3 females; mean age: 59.2 ± 15.9) were included in this study. In addition, patients that suffered from Long COVID/PASC (N=11; 3 males and 8 females; mean age: 55.7 ± 5.8), were included. Of these patients, three were classified with severe acute COVID-19 symptoms and they were hospitalized where they received oxygen. One person was classified as severe and on ventilation. Two patients were classified as having moderate COVID-19 symptoms, with hospitalization and oxygen. One patient was diagnosed with moderate symptoms but was not hospitalized. Four patients presented with mild acute COVID-19 symptoms and were not hospitalized. These patients suffered from persistent Long COVID/PASC symptoms for at least 2 months after they have recovered from acute COVID-19.

### Blood sample collection

Either a qualified phlebotomist or medical practitioner drew the citrated blood samples(4.5mL sodium citrate (3.2%) tubes (BD Vacutainer^®^, 369714)), via venepuncture, adhering to standard sterile protocol. Whole blood (WB) was centrifuged at 3000xg for 15 minutes at room temperature and the supernatant platelet poor plasma (PPP) samples were collected and stored in 1.5mL Eppendorf tubes at −80°C.

### Viscometry

Plasma viscosity was measured with the *micro*VISC™ viscometer (RheoSense Inc., CA United States), which uses Viscometer/Rheometer On-a-Chip (VROC^®^) microfluidic sensor technology. Prior to analysis, stored PPP aliquots from the samples were thawed from −80°C to 35°C with the use of an incubator. The samples were also kept at 35°C in the incubator during measurements to keep the temperature constant, as it one of the variables used to measure viscosity. Between each measurement, the micro-viscometer was cleaned with 1% Scienceware^®^ Aquet^®^ liquid detergent solution (Sigma-Aldrich, Z273260), in order to maintain stable viscosity measurements. Plasma viscosity was calculated according to Newton’s law of viscosity:

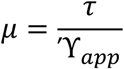

Where μ=viscosity, τ=shear stress, and Υ_app_ =apparent shear rate.

### Serum Amyloid A ELISA analysis

ELISA analysis was performed on PPP from 12 controls, 12 individuals with acute COVID-19 and 11 individuals with Long COVID/PASC. The Human SAA ELISA Kit (SAA1) (E-EL-H2183, Elabscience Biotechnology Inc.) was used in this analysis following manufacturer prescribed protocol. SAA1 is one of the two acute phase SAA proteins (Jumeau et al., 2019).

### Platelet pathology

The whole blood (WB) samples of healthy volunteers, COVID-19 and Long COVID/PASC patients were exposed to the two fluorescent markers, CD62P (PE-conjugated) (platelet surface P-selectin) (IM1759U, Beckman Coulter, Brea, CA, USA) and PAC-1 (FITC-conjugated) (340507, BD Biosciences, San Jose, CA, USA). CD62P is found in the membrane of platelets and then translocates to the platelet membrane surface. The translocation occurs after the platelet P-selectin is released from the cellular granules during platelet activation (Venter et al., 2020, Grobler et al., 2020). 4µL CD62P and PAC-1 was added to 20µL WB. The WB exposed to the markers was incubated for 30 minutes (protected from light) at room temperature. The excitation wavelength for PAC-1 was set at 450 to 488nm and the emission at 499 to 529nm and for the CD62P marker it was 540nm to 570nm and the emission 577nm to 607nm. Processed samples were viewed using the Zeiss Axio Observer 7 fluorescent microscope with a Plan-Apochromat 63x/1.4 Oil DIC M27 objective (Carl Zeiss Microscopy, Munich, Germany).

### Platelet poor plasma (PPP): Amyloid protein and anomalous clotting in platelet poor plasma samples, before and after two trypsin digestion protocols

All naïve PPP samples from T2DM, controls, acute COVID-19 and Long COVID/PASC PPP were studied using florescence microscopy. For the proteomics analysis 5 healthy, 4 Long COVID/PASC and 9 acute COVID-19 samples were used (one acute COVID-19 donor’s repeat sample, taken 2 days after the first donation were included). PPP samples were prepared for proteomics. One of the Long COVID/PASC volunteers previously donated a (healthy) blood sample. Here we also compare his healthy and Long COVID/PASC samples before and after trypsin digestion.

#### Naïve PPP samples: Fluorescence microscopy

To study anomalous clotting of fibrin(ogen) and plasma proteins, in naïve PPP samples PPP (healthy PPP, T2DM, COVID-19, Long COVID/PASC) were exposed to the fluorescent amyloid dye, Thioflavin T (ThT) (final concentration: 0,005mM) (Sigma-Aldrich, St. Louis, MO, USA) for 30 minutes at room temperature. After incubation, 4uL PPP and placed on a glass slide and covered with a coverslip. The excitation wavelength for ThT was set at 450nm to 488nm and the emission at 499nm to 529nm and processed samples were viewed using a Zeiss Axio Observer 7 fluorescent microscope with a Plan-Apochromat 63x/1.4 Oil DIC M27 objective (Carl Zeiss Microscopy, Munich, Germany) (Grobbelaar et al., 2021, Pretorius et al., 2020, Venter et al., 2020).

#### Two trypsin digestions protocols of platelet poor plasma (PPP)

Two trypsin digestion protocols were followed (See Figure 1). The first trypsin digestion protocol was followed using PPP from 5 healthy, 4 Long COVID/PASC and 9 acute COVID volunteers (one acute COVID-19 donor’s repeat sample, taken 2 days after the first donation was included). The supernatant proteomics data was collected from these samples. The trypsin digestion was also done with 4 volunteers with T2DM. Here we also included a repeat sample of one of the volunteers to confirm results. The second-step trypsinization protocol was performed on the undigested pellet deposits from 5 acute COVID-19 samples and 4 Long COVID/PASC samples. This second-step trypsinization was also performed on 5 control plasma samples.

**Figure 1:**
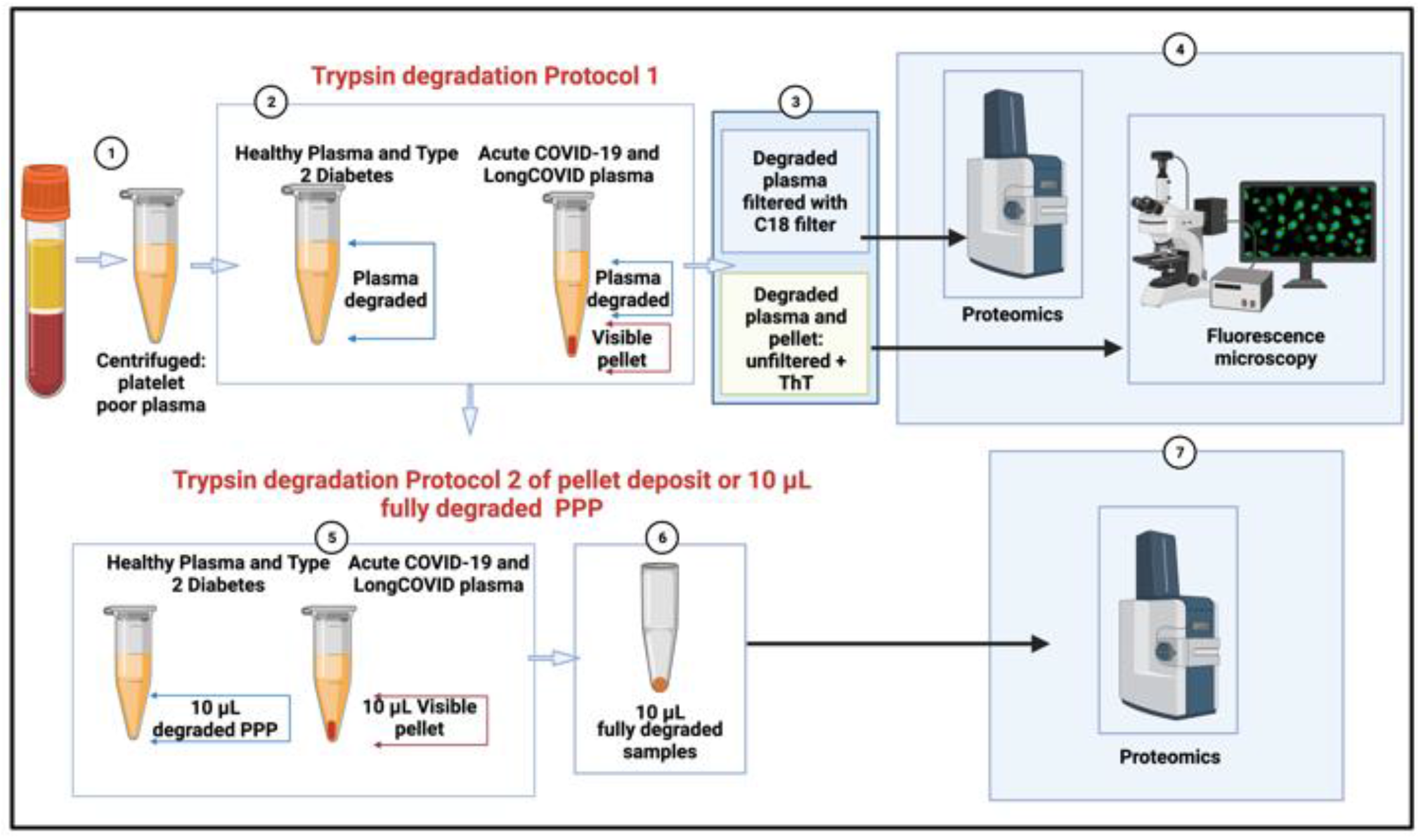
Two trypsin digestion protocols, followed by fluorescence microscopy and proteomics of platelet poor plasma (PPP) from healthy individuals, patients with type 2 diabetes Mellitus (T2DM), COVID-19 and Long COVID/PASC. **(1)** Citrated blood was centrifuged to obtain PPP. (**2)** PPP were treated with trypsin to allow plasma protein digestion. Health PPP and T2DM PPP were fully degraded. COVID-19 and Long COVID/PASC sample formed a undigested pellet deposit at the bottom of the tubes. **(3 and 4)** For fluorescence microscopy, the supernatants were removed and the remaining 10 µL of supernatant and/or pellet samples were exposed to thioflavin T (ThT) and viewed with fluorescence microscope. Before liquid chromatography-mass spectrometry (LC-MS) based proteomics, supernatants were passed through a C_18_ solid phase extraction (SPE) device. **(5)** A second trypsin digestion protocol was followed to **(6)** degrade the pellet deposit in the COVID-19 and Long COVID/PASC samples. The same method was followed with healthy and T2DM PPP (although these samples did not contain a visible pellet deposit). **(7)** Double-trypsinized samples from controls, COVID-19 and Long COVID/PASC samples were then studies using proteomics. (Figure created with BioRender.com).

##### First trypsin digestion protocol of naïve PPP

All PPP samples were exposed to a first step of trypsin digestion of the plasma proteins. The samples were diluted in 10 times in 10mM ammonium bicarbonate and protein concentration determined. The samples were standardised to 1mg/mL total protein. A total of 1µg trypsin (New England Biosystems) was added to the plasma for 1:50 enzyme to substrate ratio. No reduction or alkylation was performed. After this first trypsin digestion protocol, both COVID-19 and Long COVID/PASC samples formed a visible pellet deposit at the bottom of the tubes after centrifugation for 30 minutes at 13 000x*g*. Healthy PPP samples and the T2DM PPP samples did not form any visible deposit at the bottom of the tube.

##### Fluorescence microscopy of trypsin-degraded supernatant and visible pellet deposits

After the first trypsin digestion protocol, supernatants were removed and the remaining 10 µL was exposed to ThT (as previously described) and the rest of the supernatants were analysed using mass spectrometry (seen methods below).The 10 µL of healthy PPP and T2DM PPP contained no pellet deposit, while the 10µL of the COVID-19 and Long COVID/PASC sample did contain a visible pellet deposit. These samples were also visualized using the Zeiss Axio Observer 7 fluorescent microscope with a Plan-Apochromat 63x/1.4 Oil DIC M27 objective (Carl Zeiss Microscopy, Munich, Germany).

#### Pellet deposit digestion by a second trypsin digestion protocol

60µL of chloroform was added to the remaining 10 µL PPP, containing the pellet deposit and again centrifuged at 13 000xg for 30 min. 50µL supernatant was removed and the rest airdried. This deposit was further dissolved in 100mM Tris (pH 8.5) containing 1% (sodium dodecyl sulphate (SDS) and 5mM (tris(2-carboxyethyl)phosphine) (TCEP, Sigma), and reduced at 45°C for 1 hour. The product was cooled to room temperature and Cysteine residues blocked withmethyl methanethiosulfonate (MMTS, Sigma). These samples were also studied using mass spectrometry. The reduced and thiomethylated protein samples were diluted 1:1 with 200mM ammonium acetate (Sigma) containing 30% acetonitrile (ACN,Romil), pH 4.5. The samples were incubated with HILIC functionalised magnetic particles (ResynBiosciences) equilibrated with 100mM ammonium acetate containing 15% ACN, pH 4.5 for 30 minutes. After, binding, the supernatant was removed and the particles washed twice with 95% ACN. To each sample 0.1mg of trypsin was added in 10 mM ammonium bicarbonate. The samples were incubated over night at 37°C with agitation. After 18 hours the supernatant was removed and the particles washed with 1% trifluoroacetic acid (TFA, Sigma). The wash was combined with the first supernatant and applied to a C_18_ SPE device prior to analysis.

### Proteomics

After the first trypsin digestion protocol, the supernatants were subjected to C_18_ and solid phase extraction (SPE) and proteomics were performed. After the pellet deposit was solubilized in the second trypsin digestion protocol, the now soluble pellet deposits were also studied using proteomics.

#### Liquid chromatography on degraded supernatant and degraded pellet deposit

##### Dionex nano-RSLC

Liquid chromatography was performed on a Thermo Scientific Ultimate 3000 RSLC equipped with a 20mm x 100µm C_18_ trap column (Thermo Scientific) and a CSH 25cmx75µm 1.7µm particle size C_18_ column (Waters) analytical column. The solvent system employed was loading: 2% acetonitrile:water; 0.1% FA; Solvent A: 2% acetonitrile:water; 0.1% FA and Solvent B: 100% acetonitrile:water. The samples were loaded onto the trap column using loading solvent at a flow rate of 2µL/min from a temperature controlled autosampler set at 7°C. Loading was performed for 5 minutes before the sample was eluted onto the analytical column. Flow rate was set to 300nL/minute and the gradient generated as follows: 5.0% −30%B over 60 minutes and 30-50%B from 60-80 minutes. Chromatography was performed at 45°C and the outflow delivered to the mass spectrometer.

#### Mass spectrometry

Mass spectrometry was performed using a Thermo Scientific Fusion mass spectrometer equipped with a Nanospray Flex ionization source. Plasma samples, before and after addition of spike protein addition (1 ng.mL^-1^ final exposure concentration), from4 of our control samples were analysed with this method. The sample was introduced through a stainless-steel nano-bore emitter Data was collected in positive mode with spray voltage set to 1.8kV and ion transfer capillary set to 275°C. Spectra were internally calibrated using polysiloxane ions at m/z = 445.12003. MS1 scans were performed using the orbitrap detector set at 120 000 resolution over the scan range 375-1500 with AGC target at 4 E5 and maximum injection time of 50ms. Data was acquired in profile mode.MS2 acquisitions were performed using monoisotopic precursor selection for ion with charges +2-+7 with error tolerance set to +/- 10ppm. Precursor ions were excluded from fragmentation once for a period of 60s. Precursor ions were selected for fragmentation in HCD mode using the quadrupole mass analyser with HCD energy set to 30%. Fragment ions were detected in the Orbitrap mass analyzer set to 30 000 resolution. The AGC target was set to 5E4 and the maximum injection time to 100ms. The data was acquired in centroid mode.

#### Mass spectrometry data analysis

The raw files generated by the mass spectrometer were imported into Proteome Discoverer v1.4 (Thermo Scientific) and processed using the Sequest HT algorithm. Database interrogation was performed against the 2019-nCOVpFASTA database. Semi-tryptic cleavage with 2 missed cleavages was allowed for. Precursor mass tolerance was set to 10ppm and fragment mass tolerance set to 0.02 Da. Demamidation (NQ), oxidation (M) allowed as dynamic modifications. Peptide validation was performed using the Target-Decoy PSM validator node. The search results were imported into Scaffold Q+ for further validation (www.proteomesoftware.com) and statistical testing. A t-test was performed on the datasets and the total spectra quantitative method used to compare the datasets.

### Statistics

Statistical analysis was done using Graphpad Prism 8 (version 8.4.3). All data were subjected to Shapiro-Wilks normality tests. An unpaired T-test was performed on parametric data with the data expressed as mean ± standard deviation, whereas the Mann-Whitney U test was used on unpaired non-parametric data and the data expressed as median [Q1 – Q3] (all two-tailed).

### Supplementary material and raw data

All supplementary material and raw data can be accessed here: https://1drv.ms/u/s!AgoCOmY3bkKHi4M_1rPgXqXoq1XXSw?e=Uq0at7

## RESULTS

### Viscometry

In the current analysis, we analysed PPP from 13 controls, 10 T2DM, 13 acute COVID-19 and 11 Long COVID/PASC patients. Data was normally distributed and unpaired T-tests showed that there were no differences in PPP viscosity of controls and T2DM (p = 0.3), and that of controls and Long COVID/PASC PPP samples (p = 0.9). A significant difference was noted between PPP viscosity of controls and acute COVID-19 (p = 0.001) and acute COVID-19 and Long COVID/PASC (p = 0.002). These results suggest that the viscosity of PPP from patients with acute COVID-19 were the only samples showed an increase plasma viscosity.

### Serum Amyloid A ELISA analysis

In the current analysis, we analysed PPP from 12 controls, 12 acute COVID-19 and 11 Long COVID/PASC patients. We were specifically interested in SAA1 concentrations in plasma. The human SAA protein family comprises the acute phase SAA1/SAA2, known to activate a large set of innate and adaptive immune cells, and the constitutive SAA4 (Jumeau et al., 2019). ELISA analysis of SAA (1 to 4) is usually performed using serum samples. Serum SAA concentrations have recently been shown to be significantly and positively associated with higher COVID-19 severity and mortality (Zinellu et al., 2021). For a comprehensive review on SAA, see (Sack, 2018). Reports of serum SAA concentrations in COVID-19 patients are between 10 and 300 mg.L^-1^ (SAA type not indicated) (Cheng et al., 2020); and can be over 200 mg.L^-1^ for SAA1 (Li et al., 2020b). We analysed our data with the Mann-Whitney test. The concentration distribution of SAA1 was controls (2.8 mg.L^-1^ [2.4 - 3.8]); acute COVID-19 (3.6 mg.L^-1^ [3.3 - 5.1]) and Long COVID/PASC (median: 3.8 mg.L^-1^ [3.5 - 4.2]). There was a significant difference in PPP SAA1 concentrations between controls and acute COVID-19 (p = 0.02), and controls and Long COVID/PASC (p = 0.009). There were no significant differences between PPP SAA1 concentrations acute COVID-19 and Long COVID/PASC (p=0.45).

### Platelet pathology

Previously, we (and others) noted that platelets are hyperactivated in T2DM. Recently, we also confirmed this observation in COVID-19 samples (Venter et al., 2020). Here we show that, in the current sample, platelets from Long COVID/PASC patients are also hyperactivated (Fig. 2E and F), with an addition feature of clumped together platelets (Fig. 2G and H) not seen in COVID-19 patients.

**Figure 2:**
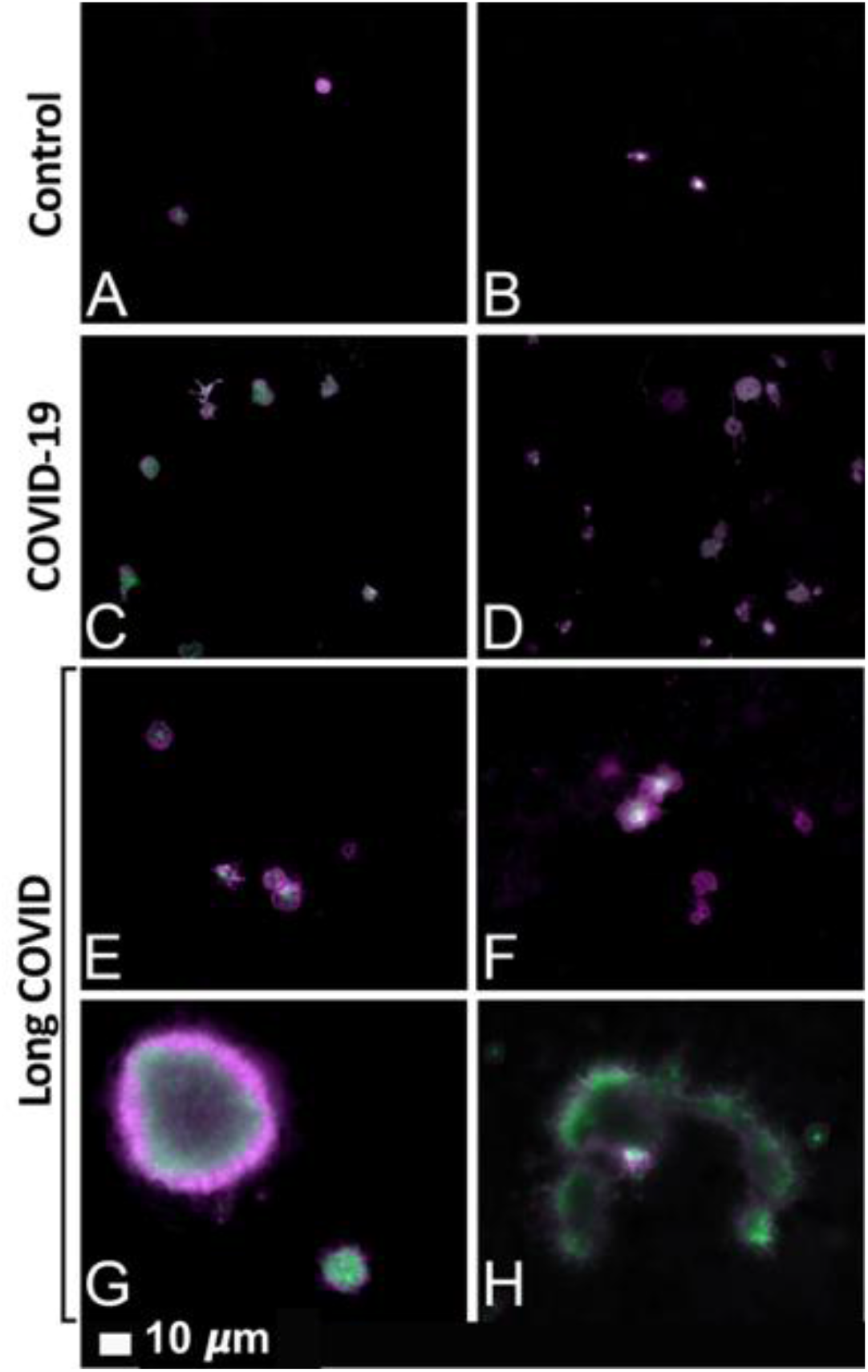
Fluorescence microscopy of whole blood samples, showing representative micrographs of hyperactivated platelets in COVID-19 **(C and D)** and Long COVID/PASC **(E-H)** samples, compared to the minimally activated control platelets **(A and B)**. Platelets were incubated with the fluorescent markers PAC-1 (green fluorescence) and CD62P-PE (purple fluorescence).

### Platelet poor plasma (PPP): Amyloid protein and anomalous clotting in platelet poor plasma samples, before and after two trypsin digestion protocols

Previously we have shown that naïve PPP (exposed to ThT) from healthy individuals and T2DM, have significantly less anomalous clotlets, compared to acute COVID-19 PPP (Pretorius et al., 2020). Here we show that PPP from Long COVID/PASC samples also have considerable anomalous (amyloid) clotlets, similar to that of acute COVID-19 PPP samples. Figure 3 shows the naïve sample of a volunteer before COVID-19 infection and during Long COVID/PASC (Figure 3A and B) and Figure 3C shows more examples of clotlets in other Long COVID/PASC patient PPP samples.

**Figure 3:**
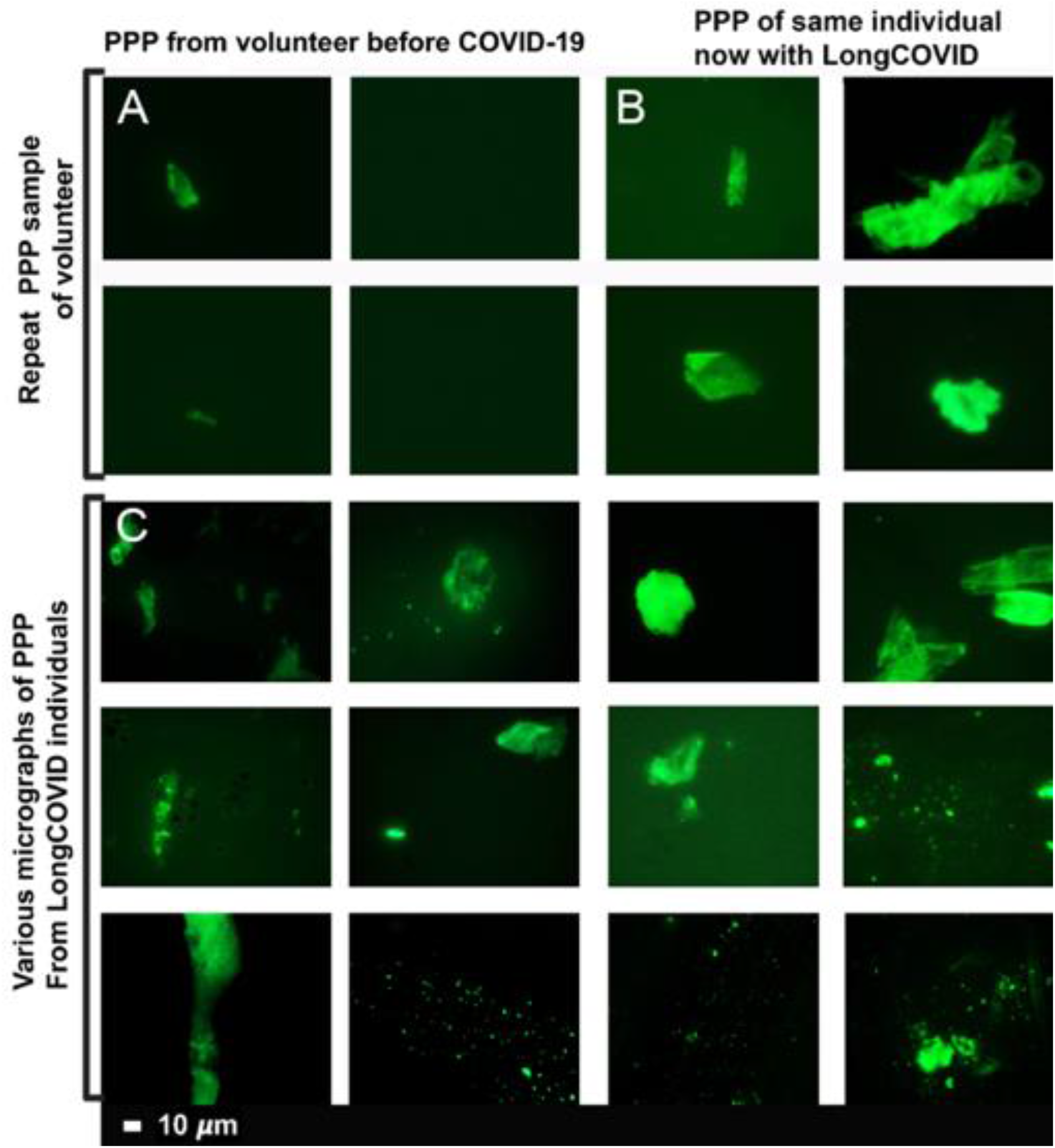
Micrographs of platelet poor plasma (PPP) (BEFORE trypsin digestion) with added thioflavin T (ThT) a healthy volunteer, before acute COVID-19 infection **(A)**, and during Long COVID/PASC **(B)**. Representative micrographs of patients with Long COVID/PASC **(C)**.

Plasma samples of healthy individuals, T2DM, COVID-19 and Long COVID/PASC patients were then exposed to the first trypsin protocol. From each sample the supernatant was removed and ThT was added to 10µL of the remaining sample and viewed with fluorescence microscopy are shown in Figure 4 and 5. In this remaining 10µL, COVID-19 and Long COVID/PASC PPP samples, a visible pellet was present.

**Figure 4:**
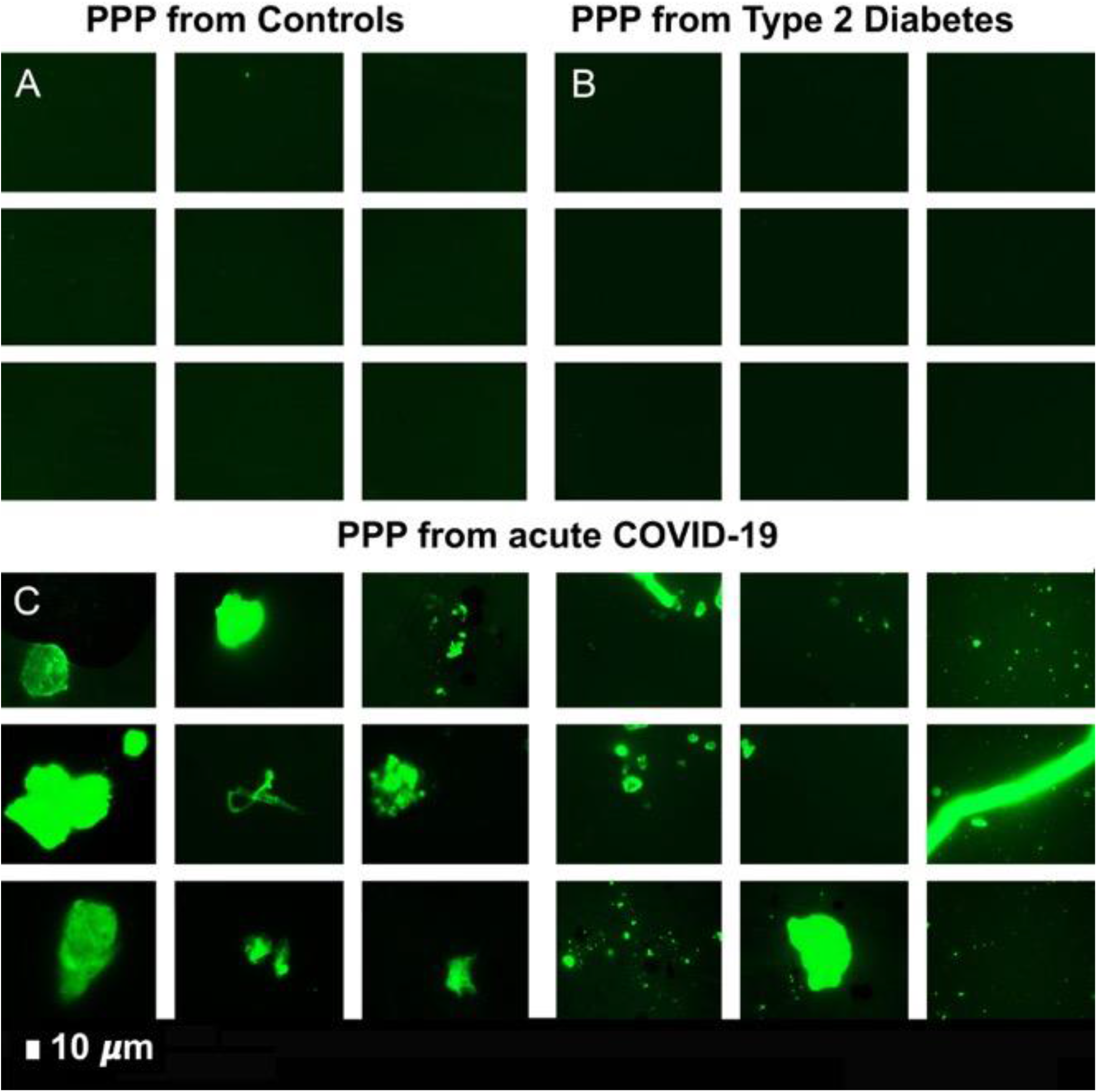
Digested supernatant of platelet poor plasma (PPP) (AFTER trypsin digestion) with added thioflavin T (ThT). **(A)** PPP from healthy individuals; **(B)** PPP from Type 2 Diabetes Mellitus **(C)** PPP from COVID-19.

**Figure 5:**
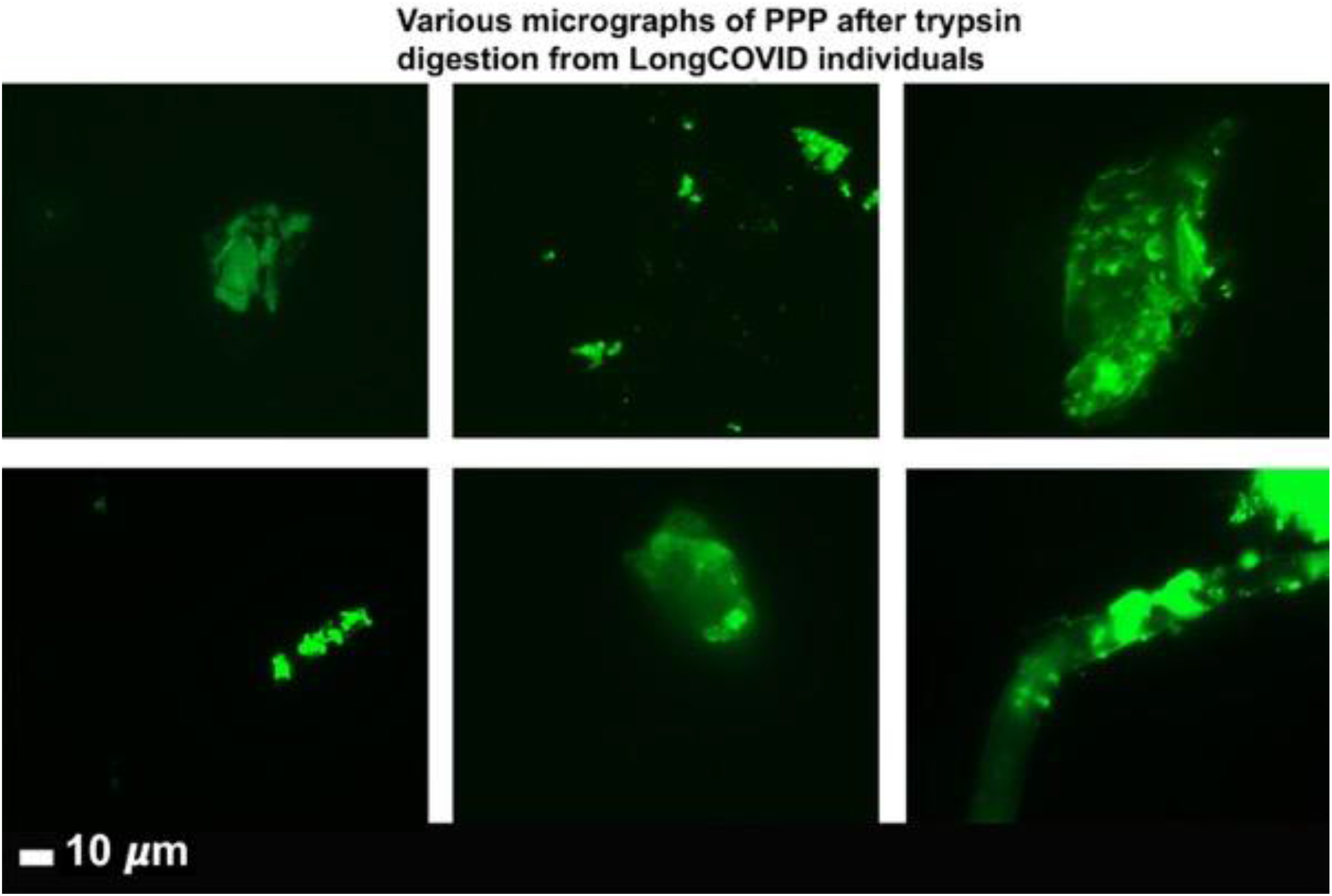
Representative micrographs of Long COVID/PASC patients. Digested supernatant (PPP) after first trypsin digestion step, where supernatant was removed and thioflavin T (ThT) added to the remaining 10 µL.

### Proteomics

In this paper we focus only on selected proteomics results of interest for clotting; e.g. fibrinogen, von Willebrand Factor (VWF), SAA4, and plasminogen and α2AP. After the first trypsinization, the PPP of the controls were fully digested, while a fibrinolytic-resistant deposit was left in the COVID-19 and Long COVID/PASC samples. Only after a second trypsinization, the pellet deposit of the COVID-19 and Long COVID/PASC could be fully digested. Proteomics were performed and our results show that various inflammatory molecules that were substantially increased in the supernatant (after first trypsinization) of COVID-19 and Long COVID/PASC samples compared to the supernatant from the controls. After the second trypsinization step, various inflammatory molecules were also substantially increased in the digested pellet deposits from the COVID-19 and Long COVID/PASC compared to the control samples. We present our results as fold changes in levels of proteins. See Table 1 for some of the most interesting results of the digested pellet deposits, as shown by fold changes (of more than 2) for the most significant proteins for pair-wise comparisons. See Figure 6 for overview plots of the protein distribution between pairwise sample comparisons (controls vs COVID-19; Controls vs Long COVID/PASC; COVID-19 vs Long COVID/PASC). All raw data for the supernatant and digested pellet data are shown in supplementary material. A striking aspect is that many proteins are increased compared to very few that are decreased.

**Table 1:** Proteomics pairwise analysis of digested pellet deposits from acute COVID-19 and Long COVID/PASC vs fully digested samples from controls. All sample types underwent a two-step trypsinization process. Controls vs acute COVID-19; Controls vs Long COVID/PASC; acute COVID-19 vs Long COVID/PASC. The proteins showed here are in both sample types; and value more than 1 = the protein more prevalent in a specific digested pellet deposit.

|  |  |  |
| --- | --- | --- |
| **Digested pellet deposits from acute COVID-19 samples vs digested plasma from Control samples** |  |  |
| These proteins are present in both sample types; and a fold change value more than 1 = the protein that more prevalent inside the digested pellet deposits from COVID-19 samples. These proteins were concentrated inside the digested pellet deposits. |  |  |
| **Protein name** | **Fold change** | **P-value** |
| Coagulation factor XIII A chain | 1.69 | 0.005 |
| von Willebrand Factor | 5.44 | 0.0001 |
| Complement component C7 | 7.84 | 0.04 |
| C-reactive protein | 15.11 | 0.0001 |
| **Digested pellet deposits from Long COVID/PASC clots samples vs digested plasma from Control samples** |  |  |
| These proteins are present in both sample types; and a fold change value more than 1 = the protein that more prevalent inside the digested pellet deposits from Long COVID/PASC samples. These proteins were concentrated inside the digested pellet deposits. |  |  |
| Plasminogen | 2.54 | 0.0004 |
| Fibrinogen alpha chain | 4.2 | 0.000001 |
| α2 antiplasmin (α2AP) | 8.89 | 0.000002 |
| von Willebrand Factor | 11.27 | 0.000001 |
| C-reactive protein | 12.19 | 0.0004 |
| Serum Amyloid A (SAA4) | 17.05 | 0.002 |
| Complement component C7 | 18.68 | 0.000002 |
| **Digested pellet deposits from Long COVID/PASC clots samples vs digested pellet deposits from acute COVID-19 samples** |  |  |
| These proteins are present in both sample types; and a fold change value more than 1 = the protein that more prevalent inside the digested pellet deposits from Long COVID/PASC samples. These proteins were concentrated inside the digested pellet deposits. |  |  |
| Plasminogen | 1.3 | 2.8 |
| Fibrinogen $\beta$ chain | 1.9 | $2.24 \times 10^{-06}$ |
| Coagulation factor XIII | 2.1 | 0.0002 |
| Fibrinogen $\alpha$ chain | 2.4 | $7.7 \times 10^{-06}$ |
| Serum Amyloid A (SAA4) | 6.5 | 0.01 |
| Complement C6 | 7.9 | 0.003 |
| α2 antiplasmin (α2AP) | 8.3 | $9.9 \times 10^{-07}$ |
| Complement factor | 24.6 | $2.2 \times 10^{-06}$ |

**Figure 6:**
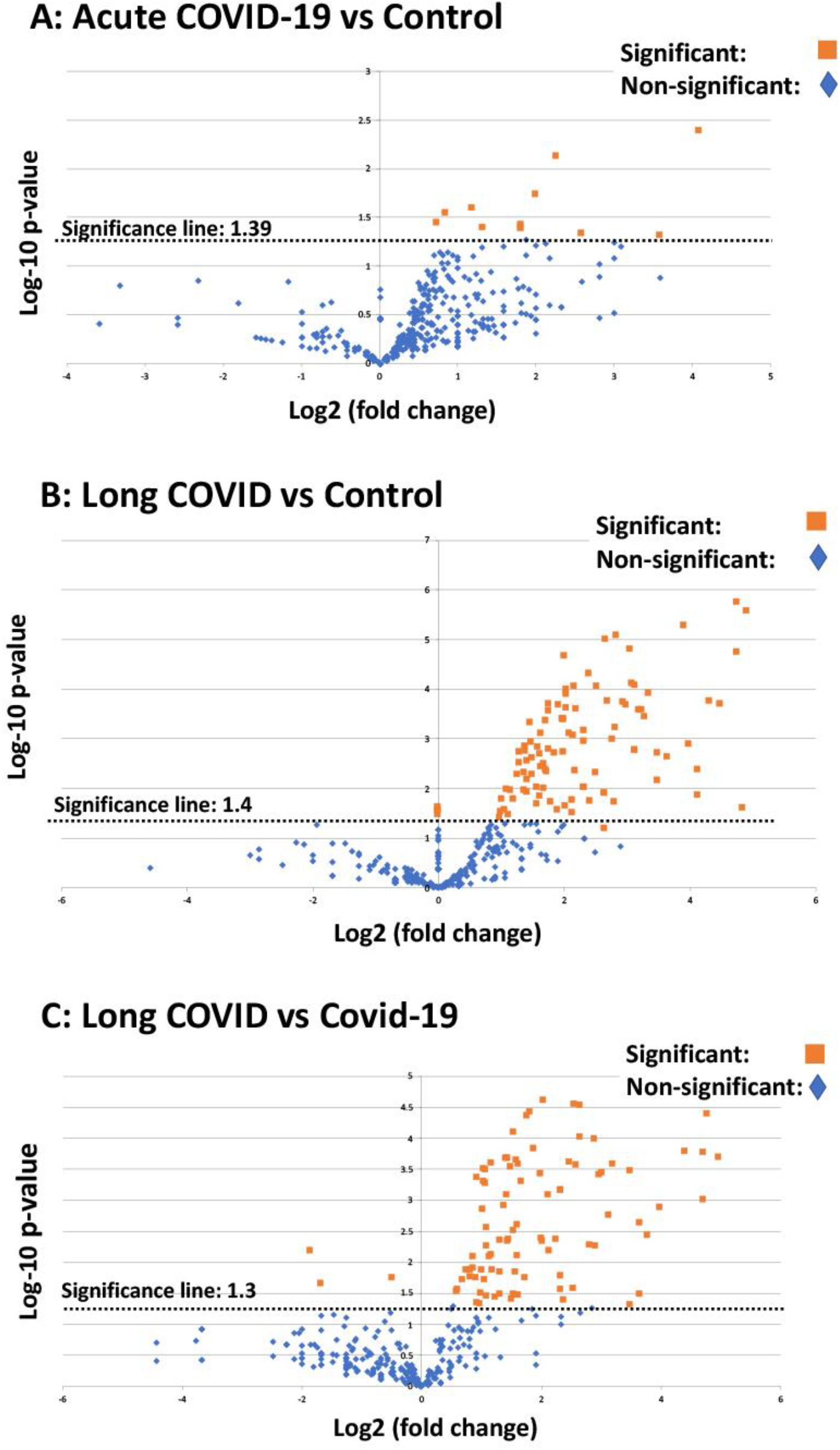
Volcano plots of the protein distribution between pairwise sample comparisons (controls vs COVID-19; controls vs Long COVID/PASC; COVID-19 vs Long COVID/PASC). All proteins above the dotted line are significant).

#### Mass spectrometry of the digested supernatants

Mass spectrometry based proteomics confirmed that the concentration of the α-fibrinogen chain is increased in the COVID-19 supernatant compared to controls and Long COVID/PASC. The γ and β chains levels were not changed in the supernatant of any of the sample types. Antiplasmin and plasminogen levels were similar in the supernatant of the controls, COVID-19 and Long COVID/PASC.

#### Mass spectrometry of the digested pellet deposits vs the digested control plasma

Mass spectrometry based proteomics confirmed that levels of the coagulation factor XII, α and β fibrinogen chains were increased in the digested pellet deposits form Long COVID/PASC samples compared to the digested plasma form controls and the digested pellet deposits from COVID-19. Relative to control and COVID-19 samples, plasminogen levels were slightly increased in Long COVID/PASC. SAA4 was not present in the digested plasma of the controls, but present in both the digested pellet deposits of samples from COVID-19 and Long COVID/PASC. It was a surprising result that SAA4 was so much increased in digested pellet deposits. There is no equivalent pellet deposit in the controls, therefore compared to the concentration of SAA4 in the digested plasma sample of the controls, presence of the concentration of SAA4 in the digested pellet deposits is interesting. We suggest that these molecules are trapped and concentrated in the fibrinolysis-resistant clots that are present in the circulation. SAA4 is a constitutively expressed molecule in contrast to SAA1 and 2 that are both acute phase proteins. It is therefore an surprizing find that SAA4 was so concentrated in the fibrinolytic-resistant pellet deposits.

## DISCUSSION

It is now well-recognized that vascular changes and thrombotic microangiopathy, diffuse intravascular coagulation and large-vessel thrombosis are major reasons for a poor COVID-19 prognosis (Ackermann et al., 2020, Iba et al., 2020). These comorbidities are linked to multisystem organ failure, as well as pulmonary vascular endothelialitis (Ackermann et al., 2020, Zhang et al., 2020). The presence of endotheliopathy in particular, is likely to be associated with critical illness and death (Goshua et al., 2020). It is also suggested that endothelial dysfunction contributes to COVID-19-associated vascular inflammation, COVID-19-associated coagulopathy, and pulmonary fibrinous microthrombi in the alveolar capillaries (Zhang et al., 2020). In some instances, patients present with a significant elevation in D-dimer/fibrin(ogen)degradation products (Wool and Miller, 2021). D-dimer and fibrin(ogen) degradation products may indicate the failing attempt of the fibrinolytic system to remove fibrin and necrotic tissue from the lung parenchyma (and also from the circulation), but being consumed or overwhelmed in the process (Medcalf et al., 2020).

Central to COVID-19 pathology is the pathological roller-coaster from hypercoagulation and hypofibrinolysis. Bouck and co-workers in 2021 found that the lag times to thrombin, plasmin, and fibrin formation were prolonged with increased disease severity in COVID-19 (Bouck et al., 2021). The authors also argue that, although the presence of D-dimer suggests fibrinolytic pathways are intact and actively dissolving (lysing) fibrin, the discovery of fibrin deposits in lungs and other organs suggests dysregulation of the balance in fibrin-forming and fibrin-dissolving (plasmin generation) pathways is a major aspect of COVID-19 pathogenesis (Bouck et al., 2021).

Results presented in the current paper point to a significant failure in the fibrinolytic process during COVID-19 and also in patients with lingering Long COVID/PASC symptoms. Our results show that plasma proteins in both COVID-19 and Long COVID/PASC plasma samples are greatly resistant to breakdown in the presence of trypsin. This was confirmed visually using fluorescence microscopy, as well as with proteomics. Most significant changes shown in proteomics analysis, were in circulating proteins related to clotting. We noted significant increases in fibrinogen chains, as well as acute phase proteins like SAA4 and α2AP, as shown in the proteomics analysis (Table 1). Our results point to a nine-fold increase in α2AP in Long COVID/PASC vs controls and an eight-fold increase in Long COVID/PASC vs acute COVID-19 for digested pellet deposits. Here we also show a difference between SAA1 and SAA4. SAA1, was about 2-fold increased in PPP from both acute COVID-19 and Long COVID/PASC as seen with an SAA1 ELISA.

SAA4 (also found as an apoliprotein of HDL), is synthesized constitutively in the liver (Sack, 2018). Here we report that SAA4 showed a significant increase in our proteomics analysis of the double trypsin-digested pellet deposits. There was a 17-fold increase in SAA4, between PPP in samples from Long COVID/PASC vs controls, and a six-fold increase when the comparison was between Long COVID/PASC over COVID-19. We report fold changes in proteins present in the fibrinolysis-resistant pellet deposits of acute COVID-19 and Long COVID/PASC, compared to the fluid sample of the controls that also underwent a double trypsinization process. It follows that the true concentrations in plasma samples may therefore not reflect the fold changes we report on here, in proteins trapped in the solubilized pellet deposits. It was recently shown that patients with low HDL-cholesterol levels at admission to the hospital were more likely to develop severe disease, compared to patients with high HDL-cholesterol levels (Feingold, 2020). Compared with the healthy controls, the patients have sharply decreased concentrations of total cholesterol, HDL-cholesterol and LDL-cholesterol (Hu et al., 2020). With reduced HDL levels in circulation, SAA4 potentially will be less partitioned into HDL.

Of particular interest is the simultaneous presence of persistent anomalous (amyloid) clotlets and a pathological fibrinolytic system. The plasmin and antiplasmin balance may be central to this phenomenon (see Figure 7). An important element of the fibrinolytic system is the conversion of circulating zymogen plasminogen to its active form plasmin (Draxler et al., 2017, Cesarman-Maus and Hajjar, 2005). Endogenous activators of plasminogen are the tissue-type plasminogen activator (tPA) and urokinase-type plasminogen activator (uPA) (Kwaan and Lindholm, 2021). The catalytic activity of tPA is largely dependent on the presence of fibrin, as both tPA and its substrate plasminogen bind to the lysine residues on fibrin, using it as a cofactor for plasmin generation (Cesarman-Maus and Hajjar, 2005). Plasmin is the effector protease of the fibrinolytic system, well known for its involvement in fibrin degradation and clot removal (Draxler et al., 2017). Plasmin is also recognized as a potent modulator of immunological processes by directly interacting with various cell types including cells of the vasculature (endothelial cells, smooth muscle cells) (Draxler et al., 2017) In fact, the removal of misfolded proteins and the maintenance of tissue homeostasis seem to be major physiological functions of plasmin (Draxler et al., 2017). Plasmin is also inhibited by the actions of various serine protease inhibitors like α2AP (Cesarman-Maus and Hajjar, 2005). High blood levels of α2AP (Singh et al., 2020), an ultrafast, covalent inhibitor of plasmin, have been linked in humans to increased risk of ischemic stroke and failure of tissue plasminogen activator therapy (Reed et al., 2017). Furthermore, plasminogen activator inhibitor-1 (PAI-1) and α2AP, maintain a delicate homeostasis in the normal physiologic state (Seheult et al., 2020). α2AP is covalently cross-linked to fibrin in the thrombus by activated factor XIII, a transglutaminase (Reed et al., 1991, Fraser et al., 2011) which is a major source of the resistance of *in vitro* plasma clots to plasmin-mediated fibrinolysis (Singh et al., 2020). It is therefore entirely plausible that, as we noted in our Long COVID/PASC samples, if there is both an acute or lingering overload of anomalous (amyloid) fibrin(ogen) clotlets in circulation, and 8-fold increase in α2AP, that the endogenous activators of plasminogen and the subsequent cascade of physiological events that are driven by plasmin generation, will fail. In addition, plasmin also processes the viral S-protein for its entry into the host cells (Kwaan and Lindholm, 2021), where plasmin, and other proteases cleave a newly inserted furin site in the S protein of SARS-CoV-2, extracellularly, which increases its infectivity and virulence (Medcalf et al., 2020). This involvement of plasmin in the cleaving of the viral S proteins, may also further contribute to a decreased efficiency of plasmin to act on an increase in anomalous fibrin(ogen) load.

**Figure 7:**
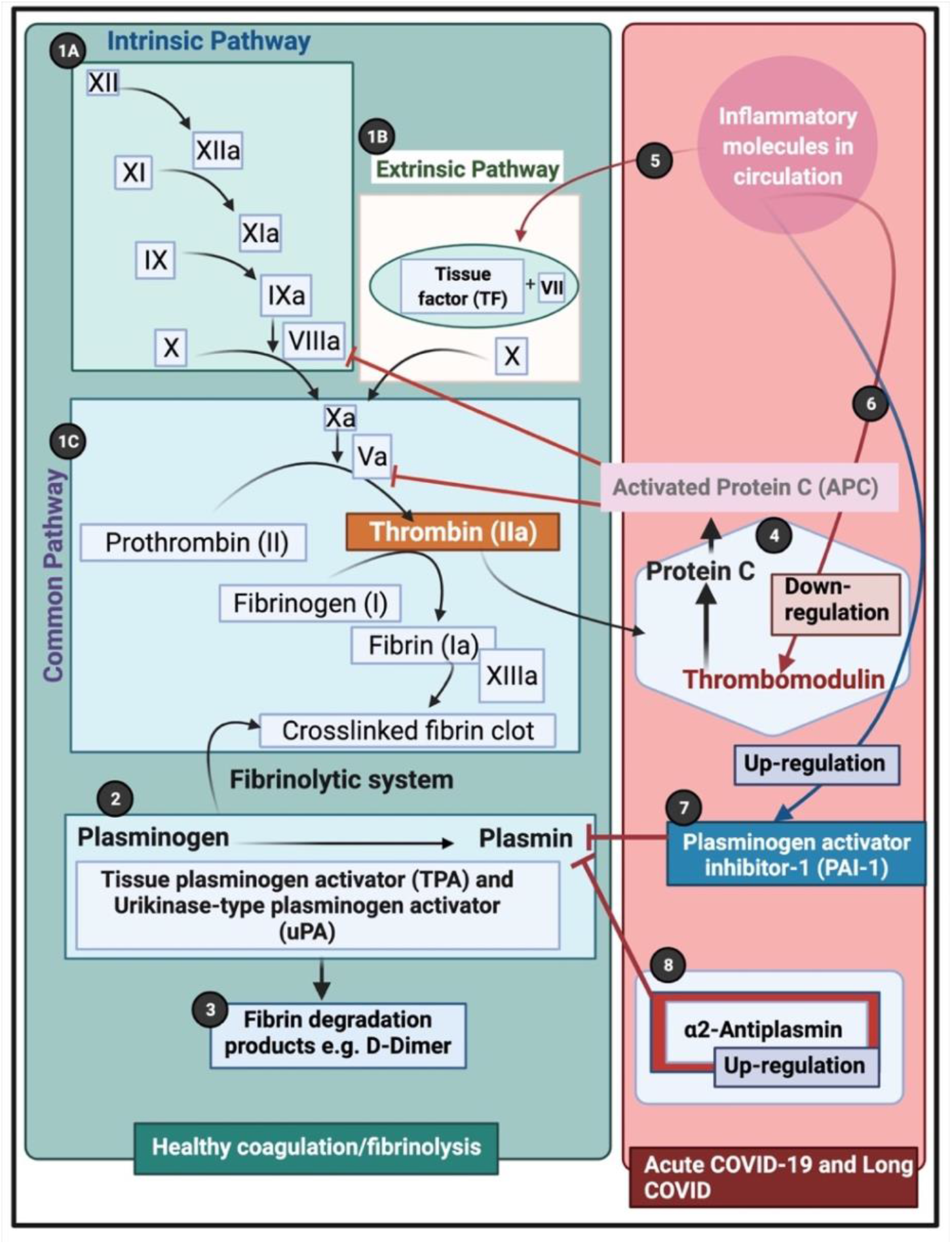
Simplified coagulation diagram (adapted from (Kell and Pretorius, 2015, de Waal et al., 2021, Miszta et al., 2021) depicting healthy and pathological processes. (**1A)** The intrinsic and **(1B)** extrinsic pathways converge into the **(1C)** common pathway. These pathways lead to the conversion of soluble fibrinogen to insoluble fibrin, catalysed by thrombin. **(2)** Tissue plasminogen activator (tPA) or urokinase-type plasminogen activator (uPA) converts plasminogen into plasmin. A healthy fibrinolytic system regulates the coagulation pathway and assists with successful lysis of the insoluble fibrin clot. **(3)** Plasmin cleaves fibrin into fibrin degradation products (FDPs), including D-dimer. **(4)** Protein C and thrombomodulin both regulate coagulation: thrombin binds to its receptor, thrombomodulin, resulting in activated protein C (APC). APC then inhibits both Va and VIIIa. **(5)** Dysregulated inflammatory molecules may interfere with tissue factor (TF) expression. **(6)** Dysregulated inflammatory molecules may also down-regulate thrombomodulin, resulting in hypercoagulation, as Va and VIIIa activities are then not sufficiently modulated. **(7)** Dysregulated inflammatory molecules in circulation can inhibit of the fibrinolytic system via up-regulation of plasminogen activator inhibitor-1 (PAI-1). PAI-I upregulation interferes with tissue plasminogen activator (TPA) function, and ultimately results in a dysregulated coagulation system. **(8)** α2-antiplasmin (α2AP) inhibits plasmin and ultimately will prevent sufficient fibrinolysis to happen. (Figure created with Biorender.com)

## CONCLUSION

Hypercoagulability is an increasingly recognized complication of COVID-19 infection, and anticoagulation has become central in the comprehensive COVID-19 management (Thachil et al., 2020, Kwaan and Lindholm, 2021). Significant anomalous (amyloid) clotlet formation that are resistant to fibrinolysis, increased α2AP and the surge of acute phase inflammatory molecules, may therefore be central contributors in the multiple coagulation/fibrinolysis pathophysiology of both COVID-19 infection and its lingering phenotype, Long COVID/PASC. We recognize that some of the proteomics findings should now be confirmed using a larger sample set as the statistical power of the current set is limited. Here we conclude that (i) hypercoagulability due to significant increases in inflammatory molecules, (ii) circulating clotlets and hyperactivated platelets, and (iii) an aberrant fibrinolytic system, are all driven by a dysfunction in clotting protein and lytic enzyme supply and demand. Central to hypofibrinolysis and persistent clotlets is the presence of a significant increase in α2AP. Clotting pathologies in both acute COVID-19 infection and in Long COVID/PASC might therefore benefit from a following a regime of continued anticlotting therapy to support the fibrinolytic system function.

## Data Availability

A data availability link is provided in the paper.

## DECLARATIONS AND AXKNOWLEDGEMENTS

We wish to thank Simone Turner and Mireille Grobler for their assistance with the ELISA assay and assistance with the curation of the samples. We also wish to thank Dr Amy Proal from PolyBio Research Foundation for comments on the paper.

## Consent for publication

All authors approved submission of the paper.

